# Impaired reach-to-grasp integration identifies cerebral visual impairment (CVI) in children and adults

**DOI:** 10.64898/2025.12.03.25341574

**Authors:** Candace J. Burke, Mia W. Nevin, Denver Grimm, Connor Mullin, Isabel B. Peters, Claudia C. Gonzalez, Isobel Hay, Uma Shahani, Laura Ward, Jenni M. Karl

## Abstract

**Aim:** Cerebral Visual Impairment (CVI), an underdiagnosed cause of childhood visual impairment, presents heterogeneous symptoms involving varying degrees of dorsal and ventral stream dysfunction. We investigated whether impaired reach-to-grasp integration occurs in CVI as a potential marker of dorsal stream dysfunction.

**Method:** People with CVI (children aged 7-17, n = 16; adults aged 18-25, n = 6) and control participants (children, n = 14; adults, n = 10) reached with their left hand to grasp plastic blocks with and without a blindfold. Reach-to-grasp timing and hand shaping measures were assessed using frame-by-frame video analysis.

**Results:** Even when they could see the block, people with CVI displayed prolonged grasp relative to reach durations, more static hand shaping during the reach, and increased reliance on non-visual hand shaping strategies after block contact to secure final grasp. The three measures were incorporated into a single composite Reach-to-Grasp Integration (RGI) score that distinguished people with CVI from controls.

**Interpretation:** Temporal, sensory, and functional dissociation of the reach and grasp occurs in CVI, consistent with dorsal stream dysfunction, and is captured by the RGI score. The RGI score could form the basis for CVI behavioural screening tools beyond standard visual assessments.

**What this paper adds:**

- People with CVI display impaired visually-guided integration of the reach and grasp, consistent with dorsal stream dysfunction.
- People with CVI use static preplanned hand shapes during visually-guided reaching.
- People with CVI rely on non-visual hand shaping strategies after target contact to facilitate grasping.
- The composite Reach-to-Grasp Integration (RGI) score distinguishes CVI from control children and adults.

## Introduction

Cerebral visual impairment (CVI) is a visual condition (1) with heterogeneous presentation due to brain, rather than ocular, damage (2). The dual stream theory, which distinguishes between the ventral ‘vision-for-perception’ and dorsal ‘vision-for-action’ streams, provides a framework for understanding the diverse behavioural manifestations of CVI (3–7). The ventral stream, from primary visual (V1) to inferior temporal cortex, supports conscious perception and recognition of people, objects, and surroundings (8). The dorsal stream, from V1 to posterior parietal cortex, supports unconscious processing of online visual input for guiding movements in real time (9–11). In CVI, behaviours mediated by the two streams may be affected to different degrees. People with ventral stream dysfunction may be diagnosed more readily because their symptoms – struggling to recognize faces, read text, or other aspects of conscious vision – are clearly visual in nature (2, 12–14). In contrast, people with primarily dorsal stream dysfunction struggle with tasks that require unconscious online visual guidance of movements, such as searching for objects in clutter, reaching and grasping objects, navigating stairs, or moving through complex spaces (15–20). Their symptoms are often misattributed to inattention or clumsiness, leading to misdiagnoses of attentional or cognitive disorders resulting in diagnostic overshadowing of the root cause – a deficit in unconscious online visuomotor control (2, 19). CVI diagnosis is further complicated by the fact that standard brain imaging cannot detect the neural changes underlying CVI (2), necessitating multiple costly assessments to rule out other disorders. Currently, there is no single agreed-upon screening tool to identify CVI, let alone dorsal stream dysfunction in CVI (2).

The dorsal stream enables visually-guided reach-to-grasp movements, which are supported by two partly dissociable neural networks (21–26). The dorsomedial reach network projects from superior parieto-occipital (SPOC) to dorsal premotor cortex (PMd) and primary motor (M1) cortex. It transports the hand to the target location so that the hand is positioned to complete the grasp. The dorsolateral grasp network projects from anterior intraparietal sulcus (aIPS) to ventral premotor cortex (PMv) and M1 (5, 23, 27–30). It guides the grasp; opening, shaping, and closing the hand to match the size and shape of the target to be grasped (26, 31–33). The two networks are proposed to work together, with significant modulatory input from the cerebellum (34–37), to enable skilled hand movements (5). Online visual input enables seamless integration of the reach and grasp into a single fluid movement (38). Thus, control participants visually fixate on the target as they lift their hand, first flexing their digits into a collected posture before opening, scaling, orienting and closing them to enable immediate precision gripping of the target (25) as illustrated in Figure 1, top. When vision is removed, the reach and grasp movements dissociate, each displaying distinct changes in timing, sensory guidance, and functional purpose (27, 39–41). Thus, blindfolded participants often elevate their outstretched hand during the reach, descend upon the target from above, and confirm its location by touch. Only after contact do they shape the hand, using various non-visual hand shaping strategies to extract haptic information about the target’s size, shape, and orientation to guide digit re-configuration and grasping (24, 39, 42) as illustrated in Figure 1, bottom.

**Figure 1.**
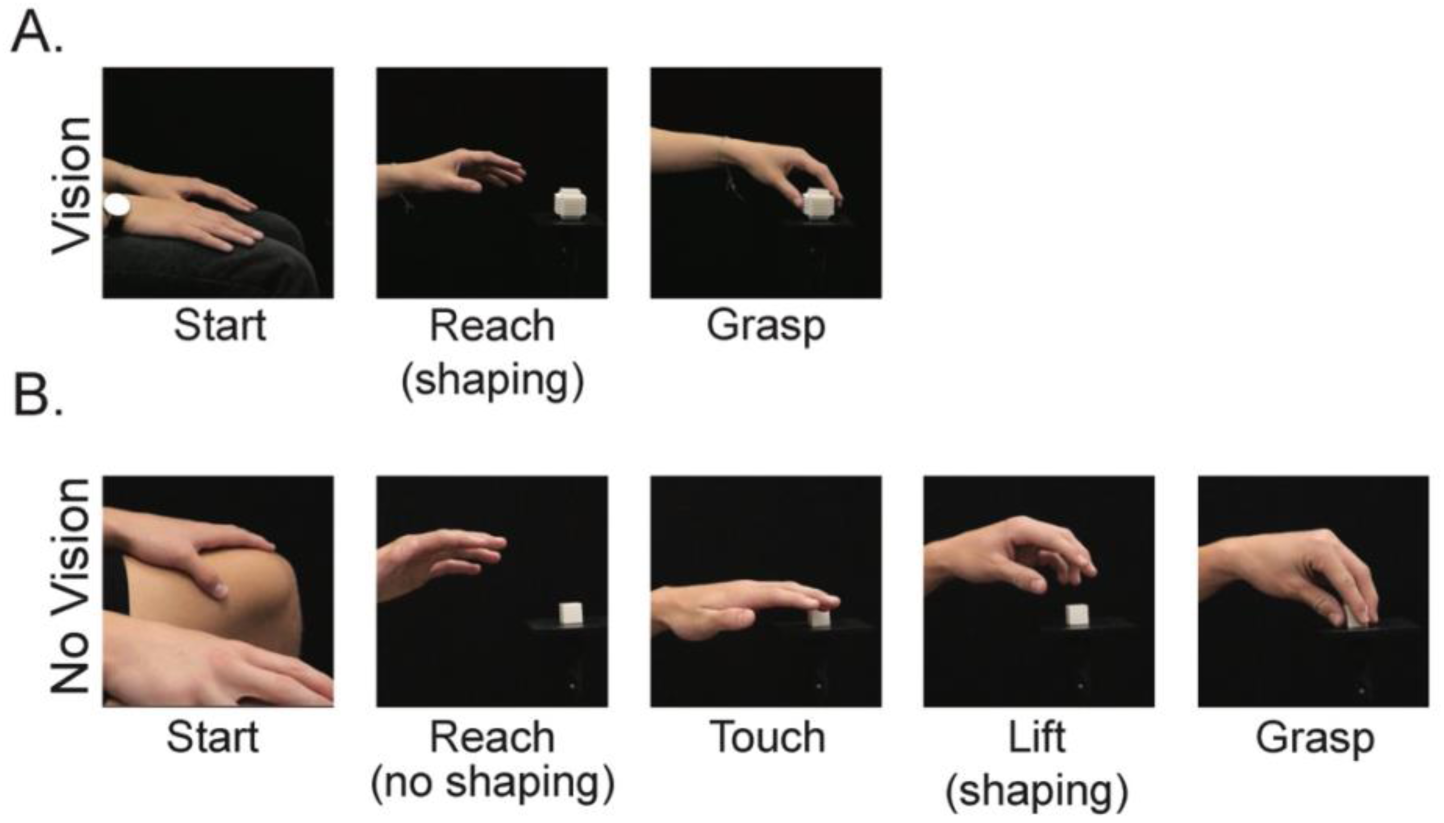
Representative still frames illustrating an integrated reach-to-grasp movement in a control participant with vision (A) and a dissociated reach-to-grasp movement in a control participant wearing a blindfold (B). Note that in the vision condition the reach and grasp are temporally integrated such that the hand preshapes and orients to the contours of the object before touching it. In contrast, the blindfolded reach and grasp are temporally dissociated such that a relatively static open and extended hand shape is maintained during the reach, which serves to locate the object by touching it, after which tactile feedback instructs hand shaping to enable grasping.

People who acquire dorsal stream dysfunction in adulthood may develop a condition called Optic Ataxia (OA) that also features dissociation of the reach and grasp (19, 43–45), despite normal visual perception (46, 47). Some people with OA can perform visually-guided reach, but not grasp, movements. They reach directly to the visible target and consistently touch it on the first attempt but do so with an open hand and only close their digits to grasp after touching it (48, 49), similar to blindfolded people. Others with OA can perform visually-guided grasp, but not reach, movements (44, 50). For example, M.H. (50) accurately opens, shapes, and closes his hand to grasp a visible target, but only if it is located immediately adjacent to his hand. If he is forced to reach for it because it is located farther away, he adopts an open-handed reach followed by a non-visually guided grasp to compensate for errors in his reach. Thus, the heterogeneous behavioural impairments observed in OA reveal that dorsal stream dysfunction can impair online visual guidance of the reach and grasp selectively, resulting in their dissociation.

Some people diagnosed with CVI also display alterations in their reach and grasp movements, suggesting dorsal stream dysfunction (19, 51), however, their impairments are typically subtle compared to OA. To investigate whether impaired visual guidance of the reach and grasp might serve as a behavioural marker of dorsal stream dysfunction in CVI, we conducted detailed frame-by-frame video analyses of CVI and control participants as they reached with and without vision to grasp plastic blocks with their left hand. We hypothesized that if the CVI participants have impaired online visual guidance of their reach and/or grasp, they should display impairments in one or both movements. Specifically, they should display greater temporal dissociation of the reach and grasp, characterized by reduced visually-guided hand shaping during the reach and greater reliance on non-visual hand shaping after object contact to enable grasping. In contrast, both groups would be expected to fully dissociate their reach and grasp movements in the blindfolded condition (39).

## Methods

### 2.1. Participants

Participants consisted of control children (n = 16; 8 male, 8 female) and children with CVI (n = 14; 8 male, 6 female) between the ages of 7 and 17; as well as control adults (n = 10; 4 male, 6 female) and adults with CVI (n = 6; 5 male, 1 female) between the ages of 18 and 25. See supplementary material for participant exclusion criteria. Participants with CVI were tested in the Vision Sciences Laboratory at Glasgow Caledonian University while control participants were tested in the Child Research Laboratory at Thompson Rivers University. All participants received a $20 gift card as compensation. The study and its procedures were approved by the Thompson Rivers University Human Research Ethics Board and the Glasgow Caledonian University Human Research Ethics Board.

### 2.2. Materials and Procedures

#### 2.2.1. Reaching targets

Participants reached with their left hand to grasp 3D printed plastic hollow blocks (Figure 2A) modelled after those by Hay et al., (19).

**Figure 2.**
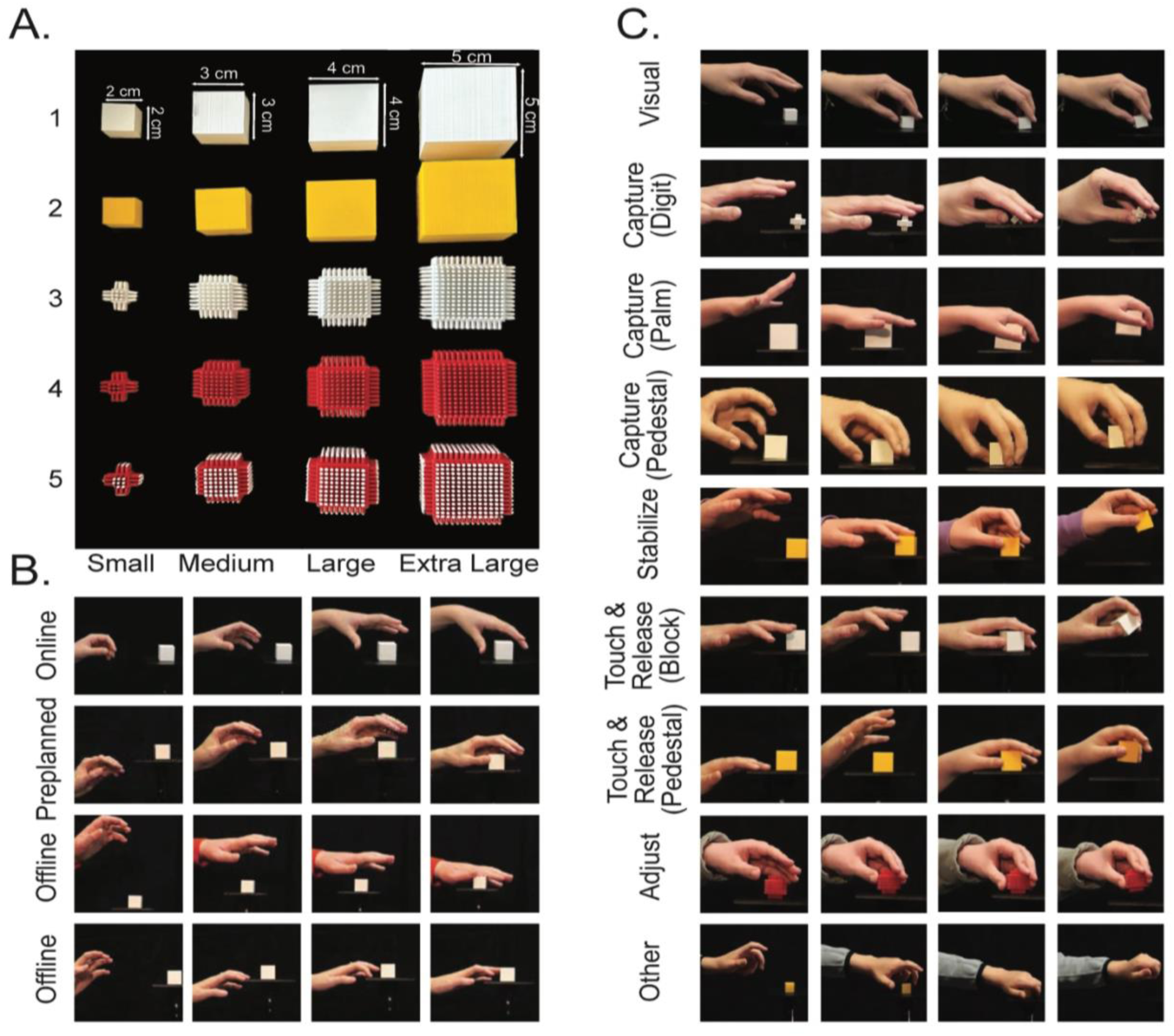
(A) Reaching targets. (B) Exemplar still frames depicting hand shaping strategies observed during the reach. (C) Representative still frames depicting hand shaping strategies observed during the grasp. Note that the online reach strategy (B, top row) and the visual grasp strategy (C, top row) are typically displayed by control participants in the vision condition and indicate intact online visual guidance of both the reach and grasp. The remaining strategies are indicative of reduced online visual guidance of hand shaping during the reach (B) and grasp (C). Note: The reaching targets were developed by NHS Dumfries & Galloway (Mountainhall Treatment Centre, Dumfries, UK) and are protected by copyright and trade secret law. All intellectual property rights remain with the copyright holder.

#### 2.2.2. Reach-to-grasp task

Participants sat in an armless chair with their palms on their thighs. A pedestal was centered at the participant’s arm length and sternum height. Two high-speed cameras (Panasonic Lumix GH6) captured frontal and midsagittal views of the participant. In the vision condition, each trial began with a “Rest” audio prompt (E-Prime 3.0 Psychology Software Tools, 2020) that cued the experimenter to place a block on the pedestal. After 15 seconds, a “Reach” prompt cued participants to reach and grasp the block with their left hand, transfer it to the experimenter, and return their hand to their thigh. Participants reached for the small blocks, from 1 to 5, followed by the medium, large, and extra-large blocks in the same order for a total of 20 reaches (Figure 2A). After an optional 5-minute break, participants repeated the task wearing a blindfold in a no vision condition.

#### 2.2.3. Movement timing and hand shaping measures

Behavioural measures, summarized in Table 1, were recorded by four trained coders using opensource software, BORIS (52). Timing measures are reported as a proportion of total reach-to-grasp movement duration (normalized time) to control for individual differences. Hand shaping measures were adapted from those reported by Karl et al. (39, 53) and divided into those that occurred before object contact (reach strategies, Figure 2B) and those that occurred after object contact (grasp strategies, Figure 2C)

**Table 1.** Timing and Hand Shaping Measures.

| Category | Measure | Definition |
| --- | --- | --- |
| Timing | Reach | Time from when the hand begins to lift from the thigh until it contacts the block |
|  | Grasp | Time from when the hand contacts the block until the grasped block lifts from the pedestal |
| Reach Strategies | Online | Smooth, efficient transitions in digit configuration involving collection, thumb-digit opposition, wrist rotation, and digit opening/closing to match block features |
|  | Preplanned | Fixed/exaggerated digit configuration or disorganized/inefficient transitions in digit configurations |
|  | Offline | Digits maintain open/extended configuration that lacks thumb-digit opposition and digit orienting to block |
| Grasp Strategies | Visual | Hand maintains a consistent grasp configuration with the digits opposed and directed to appropriate contact points enabling immediate precision grasp |
|  | Touch & Release | Initial digit contact (with block or pedestal) is released, followed by digit reorientation, reconfiguration, and one or more secondary contacts with the block before final grasp |
|  | Capture | First contact (with the digit or palm on the block or pedestal) triggers extensive closing of initially over-extended digits to achieve final grasp |
|  | Stabilize | One or more digits maintain initial stable contact with the block while the remaining digits reconfigure and reorient to grasp the block as the stabilizing digit releases |
|  | Adjust | The digits are appropriately configured but misaligned at initial contact. Initial contact is maintained while opposing digit(s) reorient to enable precision grasp |

*Note.* A small proportion (0.8%) of trials could not be categorized as any of the above grasp strategies and were coded separately as ‘Other’.

#### 2.2.4. Statistical analyses and planned comparisons

It is well established that children and adults have distinct visuomotor profiles (54, 55) and that the reach and grasp are altered when blindfolded (39, 56, 57). Accordingly, data from child and adult participants, and from the vision and no vision conditions, were analyzed separately in the Statistical Package for the Social Sciences (SPSS Version 30.0.0.0). Statistical analyses focused on a priori planned comparisons derived from the hypothesis that the CVI group would differ significantly from controls in only the vision condition. More specifically, because people with CVI are hypothesized to have a deficit in unconscious online visuomotor guidance due to dorsal stream dysfunction, we predicted that when reaching to grasp a visible target they would: 1) spend a greater proportion of total movement time transitioning from first contact to final grasp of the target (39, 57, 58), 2) display reduced online visual guidance of hand shaping during the reach as indicated by increased use of preplanned or offline reach strategies (19, 39, 53), and 3) rely more heavily on non-visual hand shaping strategies after object contact to enable the grasp (39, 59, 60). Testing planned comparisons is a standard and widely accepted approach in psychology and motor control research (61–64) that reduces the inflation of Type I error associated with multiple testing (65, 66) while also mitigating Type II error risk in studies with relatively small sample sizes (67, 68) by preventing the dilution of statistical power across comparisons outside the scope of the study (e.g., comparing controls in the vision condition to people with CVI in the no vision condition), (61–63, 69). Group differences in movement timing were analyzed using one-way analyses of variance (ANOVAs). Group differences in hand shaping measures were analyzed using mixed-effects nominal logistic regression (70) with a multinomial or binomial distribution as appropriate, a generalized logit link function, group as the fixed effect, and participant as a random effect.

## Results

### 3.1. Reach and Grasp Timing

Figure 3 illustrates that in the vision condition people with CVI spent a greater proportion of time grasping than controls did, but the two groups did not differ in the no vision condition. These results were confirmed by separate one-way ANOVAs that revealed that children, *F*(1,28) = 13.109, *p* = 0.001, *η²ₚ* = 0.319, and adults, *F*(1,14) = 8.247, *p* = .012, *η²ₚ* = 0.371, with CVI spent proportionately less time reaching and more time grasping compared to controls. In contrast, all participants spent less time reaching and more time grasping in the no vision condition, but children, *F*(1,28) = 0.274, *p* = .605, *η²ₚ* = 0.010, and adults, *F*(1,14) = 0.266, *p* = .614, *η²ₚ* = 0.019, with CVI did not differ from controls.

**Figure 3.**
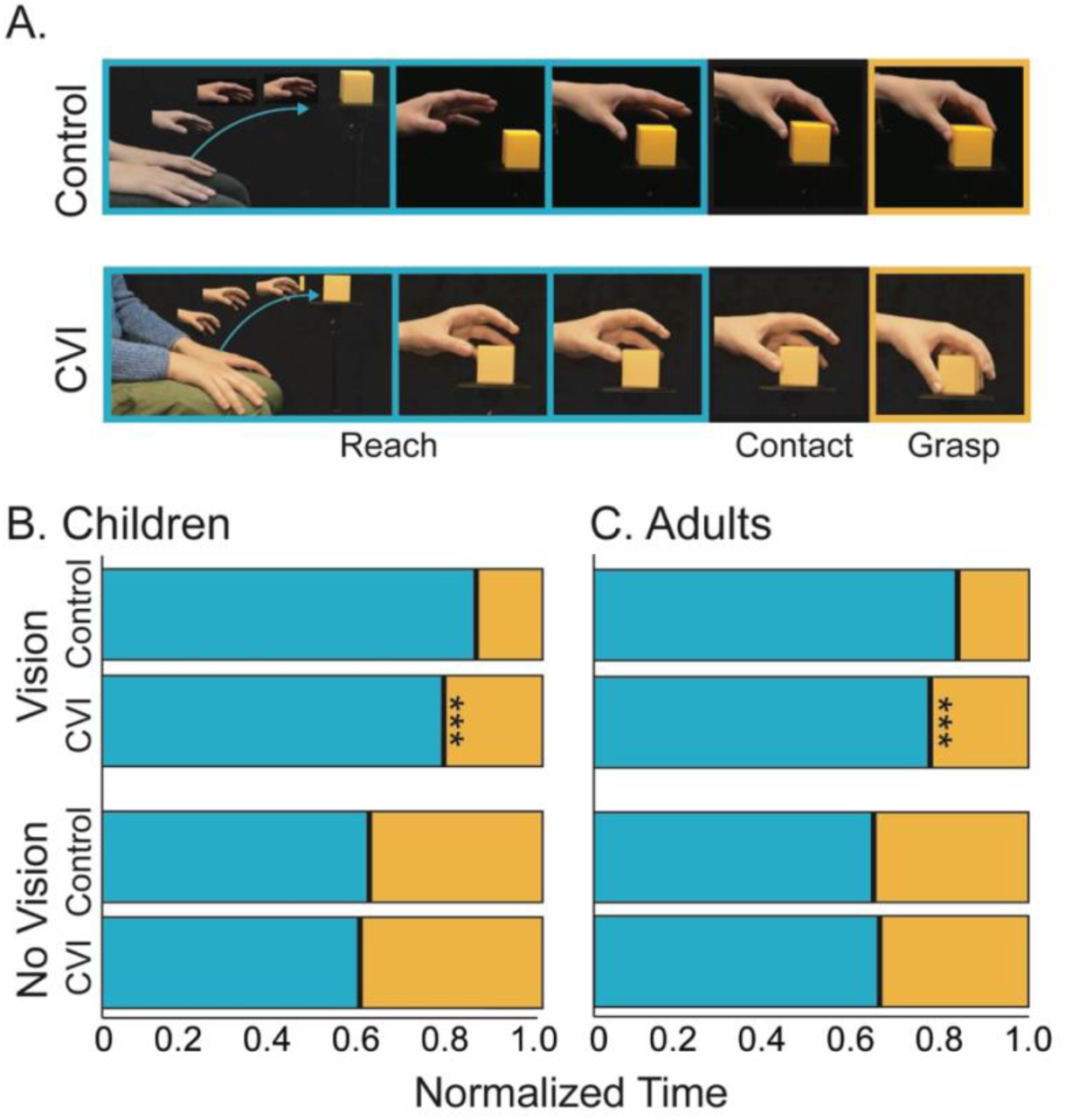
(A) Still frames depicting changes in the arm and hand’s position during the reach-to-grasp movement in a single control and CVI participant. Relative reach and grasp durations in children (B) and adults (C) with CVI compared to controls when reaching in the vision and no vision conditions. Stars indicate significant differences between the CVI and comparative control group, \*\*\**p* &<; 0.001.

### 3.2. Reach Strategies

Figures 4A&B illustrate that in the vision condition people with CVI relied more on preplanned and offline reach strategies than controls. In the no vision condition, however, the two groups behaved similarly by predominantly employing the offline reach strategy.

**Figure 4.**
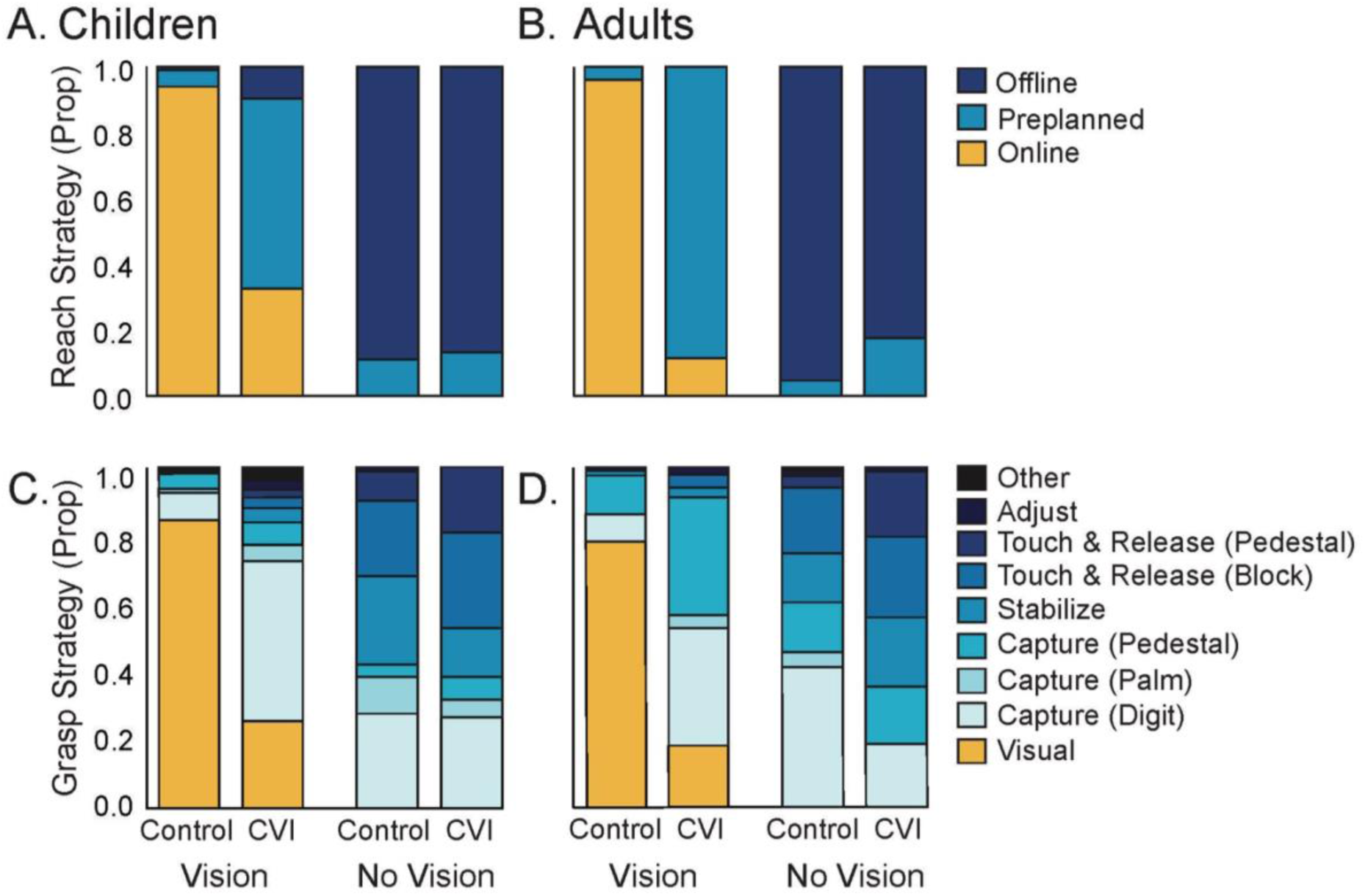
Proportional use of the different reach strategies by children (A) and adults (B) in the vision and no vision conditions. Proportional use of the different grasp strategies by children (C) and adults (D), in the vision and no vision conditions. For depictions of each strategy, see Figure 2C. Note that in the vision condition people with CVI employed the online reach strategy and the visual grasp strategy to a significantly lesser degree than their control peers.

A mixed-effects nominal logistic regression with the online strategy as the reference category revealed that group membership significantly predicted reach strategy, *F*(2, 560) = 23.04, *p* < 0.001, in children with vision. Children with CVI were significantly more likely to use an offline (*B* = 3.79, *SE* = 1.26, *OR* = 44.09, *p* = 0.003) or a preplanned (*B* = 4.07, *SE* = 0.67, *OR* = 58.34, *p* < 0.001) reach strategy compared to control children. In contrast, group membership did not significantly predict reach strategy, *F*(2, 566) = 0.14, *p* = 0.866, when the children were blindfolded. In the no vision condition neither group used the online strategy and there was no group difference in the use of the preplanned relative to offline strategy (*B* = 0.49, *SE* = 0.91, *OR* = 0.59, *p* = 0.592).

A similar pattern was observed for adults. A mixed-effects nominal logistic regression with the online strategy as the reference category revealed that group membership significantly predicted reach strategy, *F*(1, 316) = 40.15, *p* < 0.001, in adults with vision. In the vision condition, adults with CVI were significantly more likely to use the preplanned strategy (*B* = 6.03, *SE* = 0.95, *OR* = 417.28, *p* < 0.001) compared to controls and neither group used the offline strategy. In contrast, group membership did not significantly predict reach strategy, *F*(1, 318) = 0.80, *p* < 0.371, when the adults were blindfolded. In the no vision condition, neither group used the online strategy and there was no group difference in the use of the preplanned relative to offline strategy (*B* = 1.16, *SE* = 1.29, *OR* = 3.18, *p* = 0.371).

### 3.3. Grasp Strategies

Figures 4C&D illustrate that in the vision condition people with CVI relied more on non-visual grasp strategies than controls who predominantly employed a visual grasp strategy when they could see the target block. In the no vision condition, however, both groups behaved similarly, employing a variety of non-visual grasp strategies.

A mixed-effects nominal logistic regression with the visual strategy as the reference category revealed that group membership significantly predicted grasp strategy, *F*(8, 575) = 11.579, *p* < 0.001, in children with vision. Specifically, even when they could see the target, children with CVI were significantly more likely to use all the non-visual strategies except for the capture (pedestal) and touch & release (pedestal) strategies, which rarely occurred, compared to control children (see Table 2). In contrast, group membership did not significantly predict grasp strategies, *F*(7, 557) = 1.01, *p* = 0.424, in children in the no vision condition, with one exception. Children with CVI employed the touch & release (pedestal) strategy slightly more frequently than control children in the no vision condition (see Table 2).

**Table 2.** Parameter estimates for the grasp strategies.

| Condition | Grasp Strategy | B | SE | p-value | OR | Greater use in CVI than controls? |
| --- | --- | --- | --- | --- | --- | --- |
| Sighted Children | Capture (digit) | 3.10 | 0.39 | <.001*** | 22.13 | Yes |
|  | Capture (palm) | 2.35 | 1.01 | .020* | 10.52 | Yes |
|  | Capture (pedestal) | 1.03 | 0.69 | .133 | 2.81 | No reliable difference |
|  | Stabilize | 3.12 | 0.82 | <.001*** | 22.65 | Yes |
|  | Touch & release (block) | 2.47 | 0.94 | .009** | 11.85 | Yes |
|  | Touch & release (pedestal) | 18.80 | — | — | — | Unstable (rare) |
|  | Adjust | 2.68 | 0.93 | .004** | 14.61 | Yes |
| Blindfolded Children | Capture (palm) | -0.67 | 0.66 | .300 | 0.50 | No reliable difference |
|  | Capture (pedestal) | 0.19 | 0.66 | .770 | 1.21 | No reliable difference |
|  | Stabilize | -0.33 | 0.53 | .538 | 2.04 | No reliable difference |
|  | Touch & release (block) | 0.56 | 0.56 | .322 | 1.74 | No reliable difference |
|  | Touch & release (pedestal) | 1.38 | 0.68 | .042* | 3.96 | Yes |
|  | Adjust | -18.12 | — | — | — | Unstable (rare) |
| Sighted Adults | Capture (digit) | 3.24 | 0.58 | <.001*** | 25.55 | Yes |
|  | Capture (palm) | 16.98 | — | — | — | Unstable (rare) |
|  | Capture (pedestal) | 2.80 | 0.74 | <.001*** | 16.49 | Yes |
|  | Stabilize | 1.97 | 0.88 | .027* | 7.16 | Yes |
|  | Touch & release (block) | 3.34 | 1.49 | .025* | 28.31 | Yes |
|  | Touch & release (pedestal) | — | — | — | — | Never used |
|  | Adjust | 3.09 | 1.20 | 0.01** | 22.00 | Yes |
| Blindfolded Adults | Capture (palm) | -15.82 | — | — | — | Unstable (rare) |
|  | Capture (pedestal) | 0.97 | 0.74 | .190 | 2.63 | No reliable difference |
|  | Stabilize | 0.91 | 0.71 | .201 | 2.49 | No reliable difference |
|  | Touch & release (block) | 1.18 | 0.82 | .150 | 3.24 | No reliable difference |
|  | Touch & release (pedestal) | 2.04 | 1.25 | .103 | 7.65 | No reliable difference |
|  | Adjust | 0.98 | 1.52 | .522 | 2.65 | No reliable difference |

*Note.* In the vision condition, the visual strategy served as the reference category, but it was absent in the no vision condition, so the capture (digit) strategy served as the reference category.

A similar pattern was observed for adults. A mixed-effects nominal logistic regression with the visual strategy as the reference category revealed that group membership significantly predicted reach strategy, *F*(6, 306) = 8.185, *p* < 0.001, in adults with vision. Specifically, adults with CVI were significantly more likely to use every non-visual grasp strategy (except the capture (palm) and the touch & release (pedestal) strategies, which rarely or never occurred) compared to control adults (see Table 2). In contrast, group membership did not significantly predict grasp strategies in adults, *F*(6, 308) = 1.099, *p* = 0.363, when they were blindfolded in the no vision condition (see Table 2).

### 3.4. Reach-to-Grasp Integration (RGI) Score

Movement timing, reach strategy, and grasp strategy were all altered in people with CVI. Therefore, we created a single standardized Reach-to-Grasp Integration (RGI) score to capture the combined contribution of all three measures to reach-to-grasp organization. First, each measure was recoded on a 0 – 1 scale with higher values indicating greater reach-to-grasp integration:

**Reach & Grasp Timing (RGT):** Reach Duration / (Reach Duration + Grasp Duration)
**Reach Strategy (RS):** Online = 1, Preplanned = 0.5, Offline = 0
**Grasp Strategy (GS):** Visual = 1, Any Non-Visual = 0

Cronbach’s alpha for the three recoded measures was 0.859, confirming good internal consistency and reliable capture of a common underlying visuomotor construct. So then, sighted control children and adults served as reference groups to calculate three z-scores, (one for each measure: Z_RGT_, Z_RS_, and Z_GS_) for each participant. In this calculation, χ_i_ is the participant’s score, *µ_controls_* is the mean of sighted controls, and *σ_controls_*is the standard deviation of sighted controls:

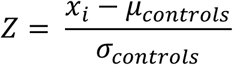

Finally, the three Z-scores were averaged to produce a single composite RGI score for each participant.

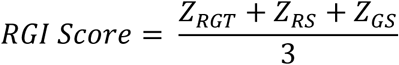

The resulting RGI scores reflect the extent to which an individual participant’s reach and grasp integration deviates from the sighted control reference group across all three measures. Scores near 0 indicate high reach and grasp integration, equivalent to sighted controls, whereas increasingly negative scores are indicative of increasingly dissociated reach and grasp movements with increased reliance on preplanned or offline hand shaping during the reach and non-visual grasp strategies after block contact.

Figure 5 illustrates that, in the vision condition, the CVI group had significantly lower RGI scores than controls, indicating that their reach and grasp movements were more dissociated even when they could see the target. These group differences disappeared in the no vision condition as all participants displayed near-to-complete dissociation of the reach and grasp when blindfolded.

**Figure 5.**
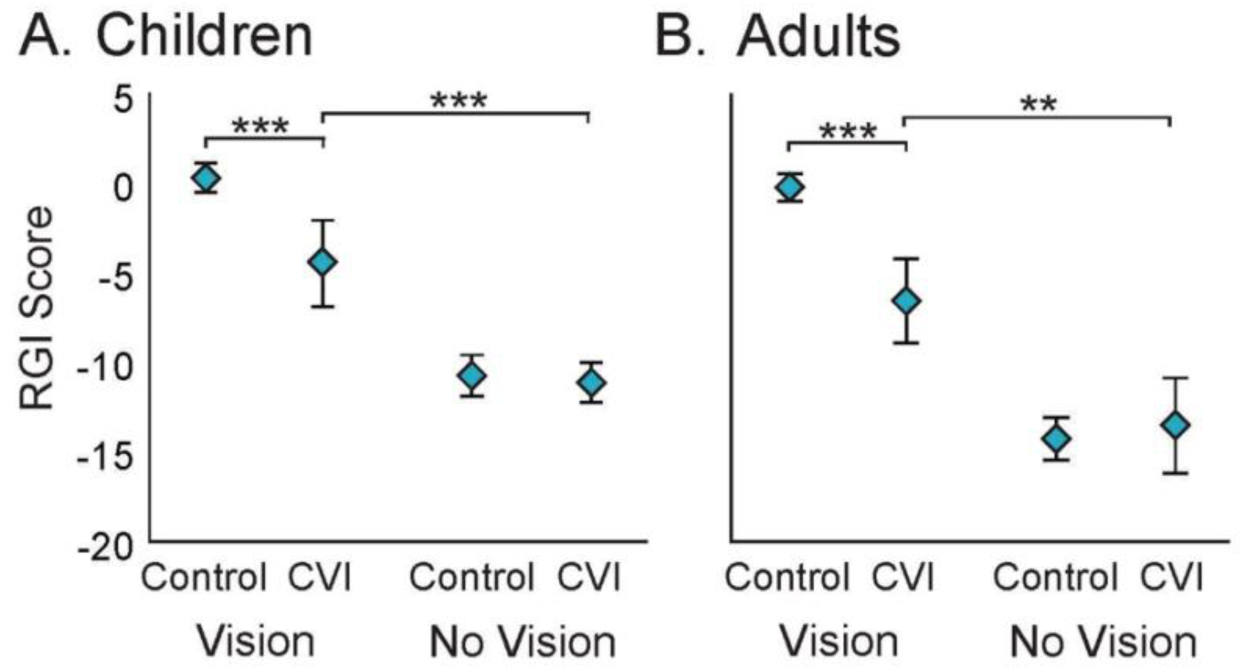
RGI scores in children (A) and adults (B) with and without CVI in the vision and no vision conditions. Stars indicate significant differences, *** *p* &<; 0.001, ** *p* &<; 0.01.

A repeated measures MANOVA was performed on RGI scores and revealed that in children there was a significant effect of group *F*(1,28) = 40.77, *p* < 0.001, *η²ₚ* = 0.593, condition *F*(1,28) = 498.88, *p* < 0.001, *η²ₚ*= 0.947, and group x condition interaction *F*(1,28) = 36.85, *p* < 0.001, *η²ₚ* = 0.568. Follow-up tests indicated that the CVI group had significantly lower RGI scores compared to controls in the vision condition, *F*(1,28) = 52.74, *p* < 0.001, *η²ₚ*= 0.653, but not the no vision condition, *F*(1,28) = 0.491, *p* = 0.489, *η²ₚ* = 0.017. Further, the RGI scores of the CVI group decreased in the no vision condition compared to the vision condition, *F*(1,13) = 75.58, *p* < 0.001, *η²ₚ* = 0.853.

In adults, there was also a significant effect of group *F*(1,14) = 26.06, *p* < 0.001, *η²ₚ*= 0.650, condition *F*(1,14) = 239.78, *p* < 0.001, *η²ₚ* = 0.945, and group x condition interaction *F*(1,14) = 26.67, *p* < 0.001, *η²ₚ* = 0.656. Follow-up tests indicated that the CVI group also had significantly lower RGI scores compared to controls in the vision condition *F*(1,14) = 64.413, *p* < 0.001, *η²ₚ* = 0.821, but not the no vision condition *F*(1,14) = 0.565, *p* = 0.465, *η²ₚ* = 0.039. Again, the RGI scores of adults with CVI decreased in the no vision condition compared to the vision condition *F*(1,5) = 19.45, *p* = 0.007, *η²ₚ* = 0.795.

## Discussion

We provide the first description of impaired visually-guided reach-to-grasp integration in children and adults with CVI (19, 51). These findings were derived from frame-by-frame video analyses of people with CVI and control participants as they reached to grasp plastic blocks with their left hand both with and without vision. Compared to controls, people with CVI spent disproportionately more time grasping, displayed largely static preplanned hand shapes during the reach, and relied more on non-visual hand shaping strategies after block contact to achieve a secure grasp even when they could see the target. These findings demonstrate that significant dissociation of the reach and grasp occurs during visually-guided prehension in CVI, consistent with dorsal stream dysfunction. In addition, by combining movement timing, reach strategy, and grasp strategy into a single Reach-to-Grasp Integration (RGI) score we generated a composite index of visuomotor function. The RGI score captures both the extent to which online visual guidance of hand shaping is impaired during the reach and whether or not compensatory reliance on other sensory modalities occurs during the grasp. It enabled robust detection of subtle deficits in unconscious visually-guided hand shaping movements in CVI and could form the basis for the future development of behavioural screening tools beyond standard visual assessments.

First, we characterized the reach-to-grasp performance of people with and without CVI both when they could see the target blocks and when they were blindfolded. Consistent with prior work (39, 40), all participants spent a much greater proportion of time grasping in the no vision condition when vision was not available to guide their movements. Importantly, CVI participants displayed a similar, albeit less pronounced, pattern even in the vision condition. Despite being able to consciously see the block, they still spent significantly more time grasping the block after contact compared to controls. This suggests that they prolong the grasp phase of the movement to compensate for an impairment in online visual guidance that normally serves to prepare the hand for grasping during the reach, prior to block contact. Still, the fact that CVI participants prolonged the grasp to an even greater extent when blindfolded suggests that they still benefitted from visual input to some extent in the vision condition.

When blindfolded, nearly all participants compensated for the complete loss of vision by using an open and extended hand to reach out and locate the block by touching it. In essence, hand shaping was almost entirely absent during the blindfolded reach. CVI participants showed a similar, but notably different, pattern even when they could consciously see the block. Instead of reaching with an open and extended hand, they adopted preplanned hand shapes that formed early and remained relatively fixed until they contacted the block. This differed from control participants who dynamically shaped their hand in preparation to grasp the object as they reached towards it. The preplanned hand shapes used by CVI participants in the vision condition resemble those of control participants when their vision is degraded but they are familiar with the features of the object they are reaching for, such as when they pantomime reaching towards known objects (71, 72), reach to remembered objects (73, 74), or reach to objects in peripheral vision (40). Under such circumstances, offline conscious visual percepts of the known object, originating in the ventral visual stream, may guide hand shaping during the reach, albeit less effectively and more effortfully, than the unconscious online visual guidance normally afforded by the dorsal stream (75–78).

Blindfolded participants also adapted to the total loss of vision by switching to non-visual grasp strategies. After touching the block, they relied on haptic feedback to adjust the position and orientation of their digits in order to securely grip the block (31, 39). Interestingly, even when they could see the block, people with CVI, but not controls, also relied on non-visual grasp strategies. Yet, their approach differed from that of blindfolded participants, who displayed more intensity and variety in their compensatory non-visual grasp strategies, employing the capture, touch & release, and stabilize strategies all relatively frequently. In contrast, when people with CVI could see the target, they strongly favoured a more modest capture strategy. Children with CVI tended to use the capture (digit) strategy while adults with CVI also employed the capture (pedestal) strategy. This consistent reliance on capture strategies, even when the block was visible, suggests that, compared to controls, people with CVI depend more on tactile feedback after contact to complete the grasp. At the same time, their reduced use of the stabilize and touch & release strategies, which are associated with more variable initial digit-to-block contact points, suggests that they are not as reliant on tactile feedback to confirm the block’s location, size, and shape, as blindfolded participants are. In other words, they likely have enough visual information from the dorsal stream to guide the reach, but not enough to fully shape the hand before contact. This may reflect functional allocation of resources within the dorsal stream, such that available online visual guidance is effectively exhausted by directing the reach movement to the correct target location, leaving hand shaping for the grasp dependent on alternate forms of guidance, such as ventral stream percepts or kinesthetic memory. Consequently, they may rely more on tactile feedback after block contact to fine-tune their grip. This interpretation aligns with neuroimaging evidence that people with CVI often exhibit reduced white matter connectivity in dorsal stream pathways (3, 15).

As movement timing, reach strategy, and grasp strategy all converged to indicate impaired online visual guidance of prehension movements in people with CVI, we developed a single measure of visuomotor performance, the composite Reach-to-Grasp Integration (RGI) score, by averaging z-transformed values for each measure. The RGI score provides a single continuous index of the extent to which the reach and grasp are temporally and functionally integrated under online visual guidance in people with CVI. It offers several unique advantages over other individual measures. From a research perspective, it reduces idiosyncratic variance from any single measure, instead emphasizing shared variance related to the underlying sensorimotor processes that support reach-to-grasp integration. Thus, it can serve as a single, robust, metric of reach-to-grasp performance in various statistical analyses, reducing complexity by eliminating the need for multiple redundant behavioural variables. Because the RGI captures shared variance across multiple measures, it should be less affected by individual differences in participants or experimental procedures. Thus, it can provide a standardized metric to compare behavioural performance across different groups of people (e.g., CVI, autism spectrum disorder (ASD), or neurotypical) and different laboratories each with their own experimental procedures. From a clinical perspective, it provides a single intuitive measure of an individual’s reach-to-grasp performance which could be used to track developmental change, performance in varied contexts, or responses to targeted interventions. Finally, given that the dorsal stream mediates the transformation of online visual input into real-time motor behaviour, the RGI could potentially serve as a behavioural proxy for dorsal stream integrity.

Some aspects of the present study require consideration and could be explored further in future research. First, although the use of blocks of varying size, colour, and texture introduced additional variability into the task, this strengthened the paradigm by minimizing the extent to which participants could rely on visual or kinesthetic memory, encouraging them to instead use online visual guidance as much as possible. Relatedly, participants always reached first with vision and then without, further reducing potential carryover effects from kinesthetic memory. Second, participants always reached with their left hand. This choice was informed by clinical reports of greater left-hand impairment in this population (Dr. Isobel Hay). Nonetheless, potential effects of both handedness and right-handed reaching should be examined in future work. Third, the RGI is standardized relative to the control group when they are reaching with vision. Thus, its interpretive value is inherently more informative for the CVI group and blindfolded controls than it is for controls with vision. Fourth, because CVI symptoms often worsen with increasing visual or environmental complexity (79, 80) future studies should examine reach-to-grasp performance in more complex visual contexts. Finally, future work should investigate the extent to which RGI scores predict other behavioural and neuroimaging measures of dorsal stream function in people with CVI.

This study is the first, to our knowledge, to characterize reach-to-grasp behaviour in both children and adults with CVI. Importantly, the results reveal that people with CVI do not simply perform as though they are completely blind. Instead, their reach-to-grasp behaviour reflects partial use of online visual guidance supported by compensatory reliance on offline conscious visual percepts, kinesthetic memory, and somatosensory feedback. These results demonstrate that CVI is fundamentally a visuomotor, rather than purely visual, disorder. By combining measures of movement timing and hand shaping during the reach and grasp, we developed the RGI score, a composite index of the effectiveness with which online visual input guides reach and grasp coordination. The RGI was highly effective at distinguishing people with CVI from both controls with vision and blindfolded participants, highlighting potential value as a behavioural marker of dorsal stream dysfunction. Beyond its research value, the RGI has strong translational potential as a behavioural screening tool for CVI with dorsal stream dysfunction. Once fully validated, it could be easily deployed in schools, vision clinics, and pediatric care settings, potentially enabling earlier identification, fewer misdiagnoses, and more targeted interventions for CVI.

## Data Availability

All data produced are available online at https://osf.io/uht5j/overview?view_only=df9b966017d245d78d4d86e6f7207da5

## Disclosure of Interests and Conflicts

The authors have stated that they had no interests which might be perceived as posing a conflict or bias and none of the authors have a conflict of interest to disclose.

## Acknowledgments

We would like to thank and acknowledge the contribution the research participants have made to this study. We would also like to thank Supreeta Ranchod for assistance with primary data collection, as well as Priya Kumar whose pilot work helped to inform methodological decisions in the current study. We thank NHS Dumfries & Galloway for permission to use their proprietary ‘Target blocks for optic ataxia diagnosis/assessment’ materials. This research was funded by the support of the Natural Sciences and Engineering Research Council of Canada (JMK) [RGPIN-2017-05995] and the Thompson Rivers University Undergraduate Research Experience Award Program (UREAP).

## Notes

### Competing Interest Statement

The authors have declared no competing interest.

### Author Declarations

The study and its procedures were approved by the Thompson Rivers University Human Research Ethics Board and the Glasgow Caledonian University Human Research Ethics Board.

## References

1. Williams C, Pease A, Warnes P, Harrison S, Pilon F, Hyvarinen L, et al. Cerebral visual impairment-related vision problems in primary school children: a cross-sectional survey. Dev Med Child Neurol. 2021;63(6):683–9.

2. Lueck AH, Dutton G. Vision and the brain: Understanding cerebral visual impairment in children: AFB Press, American Foundation for the Blind Arlington, VA; 2015.

3. Merabet LB, Mayer DL, Bauer CM, Wright D, Kran BS, editors. Disentangling how the brain is “wired” in cortical (cerebral) visual impairment. Semin Pediatr Neurol; 2017: Elsevier.

4. Reislev NL, Kupers R, Siebner HR, Ptito M, Dyrby TB. Blindness alters the microstructure of the ventral but not the dorsal visual stream. Brain Struct. Funct. 2016;221:2891–903.

5. Van Polanen V, Davare M. Interactions between dorsal and ventral streams for controlling skilled grasp. Neuropsychologia. 2015;79:186–91.

6. Vinci-Booher S, Caron B, Bullock D, James K, Pestilli F. Development of white matter tracts between and within the dorsal and ventral streams. Brain Struct. Funct. 2022;227(4):1457–77.

7. Goodale MA. Separate visual systems for perception and action: a framework for understanding cortical visual impairment. DMCN. 2013;55(s4):9–12.

8. James TW, Culham J, Humphrey GK, Milner AD, Goodale MA. Ventral occipital lesions impair object recognition but not object-directed grasping: an fMRI study. Brain. 2003;126(11):2463–75.

9. Haxby JV, Grady CL, Horwitz B, Ungerleider LG, Mishkin M, Carson RE, et al. Dissociation of object and spatial visual processing pathways in human extrastriate cortex. PNAS. 1991;88(5):1621–5.

10. Milner AD, Goodale MA. Two visual systems re-viewed. Neuropsychologia. 2008;46(3):774–85.

11. Mishkin M, Ungerleider LG. Contribution of striate inputs to the visuospatial functions of parieto-preoccipital cortex in monkeys. Behav. Brain Res. 1982;6(1):57–77.

12. Ffytche DH, Blom JD, Catani M. Disorders of visual perception. J Neurol Neurosurg Psychiatry. 2010;81(11):1280–7.

13. Rokem A, Takemura H, Bock AS, Scherf KS, Behrmann M, Wandell BA, et al. The visual white matter: The application of diffusion MRI and fiber tractography to vision science. J. Vis. 2017;17(2):4-.

14. Ortibus ELS, Verhoeven J, Sunaert S, Casteels I, De Cock P, Lagae L. Integrity of the inferior longitudinal fasciculus and impaired object recognition in children: a diffusion tensor imaging study. DMCN. 2012;54(1):38–43.

15. Bauer CM, Heidary G, Koo B-B, Killiany RJ, Bex P, Merabet LB. Abnormal white matter tractography of visual pathways detected by high-angular-resolution diffusion imaging (HARDI) corresponds to visual dysfunction in cortical/cerebral visual impairment. JAAPOS. 2014;18(4):398–401.

16. Bennett CR, Bailin ES, Gottlieb TK, Bauer CM, Bex PJ, Merabet LB, editors. Virtual reality based assessment of static object visual search in ocular compared to cerebral visual impairment. 2018: Springer.

17. Dutton GN, McKillop ECA, Saidkasimova S. Visual problems as a result of brain damage in children. BMJ Publishing Group Ltd; 2006. p. 932–3.

18. Fazzi E, Bova S, Giovenzana A, Signorini S, Uggetti C, Bianchi P. Cognitive visual dysfunctions in preterm children with periventricular leukomalacia. DMCN. 2009;51(12):974–81.

19. Hay I, Dutton G, Biggar S, Ibrahim H, Assheton D. Exploratory study of dorsal visual stream dysfunction in autism; a case series. Res. Autism Spectr. Disord. 2020;69:101456.

20. McKillop E, Dutton GN. Impairment of vision in children due to damage to the brain: a practical approach. BIOJ. 2008;5.

21. Cavina-Pratesi C, Connolly JD, Monaco S, Figley TD, Milner AD, Schenk T, et al. Human neuroimaging reveals the subcomponents of grasping, reaching and pointing actions. Cortex. 2018;98:128–48.

22. Konen CS, Mruczek REB, Montoya JL, Kastner S. Functional organization of human posterior parietal cortex: grasping-and reaching-related activations relative to topographically organized cortex. J. Neurophysiol. 2013;109(12):2897–908.

23. Jeannerod M. Visuomotor channels: Their integration in goal-directed prehension. Hum. Mov. Sci. 1999;18(2-3):201–18.

24. Karl JM, Schneider LR, Whishaw IQ. Nonvisual learning of intrinsic object properties in a reaching task dissociates grasp from reach. Exp. Brain Res. 2013;225:465–77.

25. Karl JM, Wilson AM, Bertoli ME, Shubear NS. Touch the table before the target: contact with an underlying surface may assist the development of precise visually controlled reach and grasp movements in human infants. Exp. Brain Res. 2018;236:2185–207.

26. Whishaw IQ, Karl JM. The contribution of the reach and the grasp to shaping brain and behaviour. CJEP. 2014;68(4):223.

27. Cavina-Pratesi C, Monaco S, Fattori P, Galletti C, McAdam TD, Quinlan DJ, et al. Functional magnetic resonance imaging reveals the neural substrates of arm transport and grip formation in reach-to-grasp actions in humans. J. Neurosci. 2010;30(31):10306–23.

28. Culham JC, Valyear KF. Human parietal cortex in action. Curr. Opin. Neurobiol. 2006;16(2):205–12.

29. Filimon F. Human cortical control of hand movements: parietofrontal networks for reaching, grasping, and pointing. Neuroscientist. 2010;16(4):388–407.

30. Rizzolatti G, Luppino G. The cortical motor system. Neuron. 2001;31(6):889–901.

31. Karl JM, Whishaw IQ. Different evolutionary origins for the reach and the grasp: an explanation for dual visuomotor channels in primate parietofrontal cortex. Front. Neurol. 2013;4:208.

32. Culham J, Cavina-Pratesi C, Singhal A. The role of parietal cortex in visuomotor control: What have we learned from neuroimaging? Neuropsychologia. 2006;44:2668–84.

33. Grol MJ, Majdandzić J, Stephan KE, Verhagen L, Dijkerman HC, Bekkering H, et al. Parieto-frontal connectivity during visually guided grasping. The J. Neurosci : the official journal of the Society for Neuroscience. 2007;27(44):11877–87.

34. Marneweck M, Grafton S. Representational Neural Mapping of Dexterous Grasping Before Lifting in Humans. The J. Neurosci. 2020;40:2708–16.

35. Moulton E, Galléa C, Kemlin C, Valabregue R, Maier MA, Lindberg P, et al. Cerebello-cortical differences in effective connectivity of the dominant and non-dominant hand during a visuomotor paradigm of grip force control. Front. Hum. Neurosci. 2017;11:511.

36. Rizzo J, Beheshti M, Naeimi T, Feiz F, Fatterpekar G, Balcer L, et al. The complexity of eye-hand coordination: a perspective on cortico-cerebellar cooperation. Cerebellum Ataxias. 2020;7.

37. Zackowski KM, Thach Jr WT, Bastian AJ. Cerebellar subjects show impaired coupling of reach and grasp movements. Exp. Brain Res. 2002;146(4):511–22.

38. Allart E, Devanne H, Delval A. Contribution of transcranial magnetic stimulation in assessing parietofrontal connectivity during gesture production in healthy individuals and brain-injured patients. Neurophysiol. Clin. 2019;49(2):115–23.

39. Karl JM, Sacrey L-AR, Doan JB, Whishaw IQ. Hand shaping using hapsis resembles visually guided hand shaping. Exp. Brain Res. 2012;219:59–74.

40. Hall LA, Karl JM, Thomas BL, Whishaw IQ. Reach and Grasp reconfigurations reveal that proprioception assists reaching and hapsis assists grasping in peripheral vision. Exp. Brain Res. 2014;232(9):2807–19.

41. Whishaw IQ, Karl JM. The evolution of the hand as a tool in feeding behavior: The multiple motor channel theory of hand use. Feeding in vertebrates: Evolution, morphology, behavior, biomechanics. 2019:159–86.

42. Bozzacchi C, Brenner E, Smeets JB, Volcic R, Domini F. How removing visual information affects grasping movements. Exp. Brain Res. 2018;236:985–95.

43. Andersen RA, Andersen KN, Hwang EJ, Hauschild M. Optic ataxia: from Balint’s syndrome to the parietal reach region. Neuron. 2014;81(5):967–83.

44. Baumard J, Etcharry-Bouyx F, Chauviré V, Boussard D, Lesourd M, Remigereau C, et al. Effect of object substitution, spontaneous compensation and repetitive training on reaching movements in a patient with optic ataxia. Neuropsychol. Rehabil. 2020;30(9):1786–813.

45. Borchers S, Müller L, Synofzik M, Himmelbach M. Guidelines and quality measures for the diagnosis of optic ataxia. Front. Hum. Neurosci. 2013;7:324.

46. Pisella L, Grea H, Tilikete C, Vighetto A, Desmurget M, Rode G, et al. An ‘automatic pilot’for the hand in human posterior parietal cortex: toward reinterpreting optic ataxia. Nat. Neurosci. 2000;3(7):729–36.

47. Perenin MT, Vighetto A. Optic ataxia: A specific disruption in visuomotor mechanisms: I. Different aspects of the deficit in reaching for objects. Brain. 1988;111(3):643–74.

48. Binkofski F, Dohle C, Posse S, Stephan KM, Hefter H, Seitz RJ, et al. Human anterior intraparietal area subserves prehension: a combined lesion and functional MRI activation study. Neurology. 1998;50(5):1253–9.

49. Jeannerod M, Decety J, Michel F. Impairment of grasping movements following a bilateral posterior parietal lesion. Neuropsychologia. 1994;32(4):369–80.

50. Cavina-Pratesi C, Ietswaart M, Humphreys GW, Lestou V, Milner AD. Impaired grasping in a patient with optic ataxia: primary visuomotor deficit or secondary consequence of misreaching? Neuropsychologia. 2010;48(1):226–34.

51. Bennett CR, Bauer CM, Bailin ES, Merabet LB. Neuroplasticity in cerebral visual impairment (CVI): assessing functional vision and the neurophysiological correlates of dorsal stream dysfunction. Neurosci Biobehav Rev. 2020;108:171–81.

52. Friard O, Gamba M. BORIS: a free, versatile open-source event-logging software for video/audio coding and live observations. Methods Ecol. Evol. 2016;7(11):1325–30.

53. Karl JM, Kuntz JR, Lenhart LA, Whishaw IQ. Frame-by-Frame Video Analysis of Idiosyncratic Reach-to-Grasp Movements in Humans. JoVE. 2018(131):e56733.

54. Beck MM, Kristensen FT, Abrahamsen G, Spedden ME, Christensen MS, Lundbye-Jensen J. Distinct mechanisms for online and offline motor skill learning across human development. Dev. Sci. 2024;27(6):e13536.

55. Cook A, Aziz M, Zafar A, Giaschi D, Im HY. Developmental characteristics of visuomotor adaptation strategies in childhood. J. Vis. 2023.

56. Schettino L, Adamovich S, Poizner H. Effects of object shape and visual feedback on hand configuration during grasping. Exp. Brain Res. 2003;151:158–66.

57. Grant S, Conway M. Deficits in Reach Planning and On-Line Grasp Control in Adults With Amblyopia. Invest Ophthalmol Vis Sci. 2023;64.

58. Betti S, Castiello U, Begliomini C. Reach-to-Grasp: A Multisensory Experience. Front Psychol. 2021;12:614471.

59. Churchill A, Hopkins B, Rönnqvist L, Vogt S. Vision of the hand and environmental context in human prehension. Exp. Brain Res. 2000;134(1):81–9.

60. Rand M, Lemay M, Squire L, Shimansky Y, Stelmach G. Role of vision in aperture closure control during reach-to-grasp movements. Exp. Brain Res. 2007;181(3):447–60.

61. Furr RM, Rosenthal R. Evaluating theories efficiently: The nuts and bolts of contrast analysis. Understanding Statistics: Statistical Issues in Psychology, Education, and the Social Sciences. 2003;2(1):33–67.

62. Klockars AJ, Hancock GR. Power of a stagewise intersection protected multiple comparison procedure. Communications in Statistics-Simulation and Computation. 1996;25(4):953–60.

63. Rosenthal R, Rosnow RL, Rubin DB. Contrasts and effect sizes in behavioral research: A correlational approach: Cambridge University Press; 2000.

64. Schad DJ, Vasishth S, Hohenstein S, Kliegl R. How to capitalize on a priori contrasts in linear (mixed) models: A tutorial. Journal of memory and language. 2020;110:104038.

65. Armstrong RA. When to use the B onferroni correction. OPO. 2014;34(5):502–8.

66. Rubin M. When does HARKing hurt? Identifying when different types of undisclosed post hoc hypothesizing harm scientific progress. Rev. Gen. Psychol. 2017;21(4):308–20.

67. Maxwell SE, Delaney HD. Designing experiments and analyzing data : a model comparison perspective. 2nd ed. Mahwah, N.J.: Lawrence Erlbaum Associates; 2004.

68. Perneger TV. What’s wrong with Bonferroni adjustments. BMJ. 1998;316(7139):1236–8.

69. Abdi H, Williams LJ. Contrast analysis. Encyclopedia of research design. 2010;1:243–51.

70. Hedeker D. A mixed-effects multinomial logistic regression model. Stat Med. 2003;22(9):1433–46.

71. Kuntz JR, Whishaw IQ. Synchrony of the Reach and the Grasp in pantomime reach-to-grasp. Exp Brain Res. 2016;234(11):3291–303.

72. Fukui T, Inui T. How vision affects kinematic properties of pantomimed prehension movements. Front Psychol. 2013;4:44.

73. Cornelsen S, Rennig J, Himmelbach M. Memory-guided reaching in a patient with visual hemiagnosia. Cortex. 2016;79:32–41.

74. Hesse C, Franz VH. Memory mechanisms in grasping. Neuropsychologia. 2009;47(6):1532–45.

75. Cohen NR, Cross ES, Tunik E, Grafton ST, Culham JC. Ventral and dorsal stream contributions to the online control of immediate and delayed grasping: a TMS approach. Neuropsychologia. 2009;47(6):1553–62.

76. Goodale MA. Lessons from human vision for robotic design. Autonomous Intelligent Systems. 2021;1(1):2.

77. Monaco S, Gallivan JP, Figley TD, Singhal A, Culham JC. Recruitment of foveal retinotopic cortex during haptic exploration of shapes and actions in the dark. J. Neurosci. 2017;37(48):11572–91.

78. Singhal A, Monaco S, Kaufman LD, Culham JC. Human fMRI reveals that delayed action re-recruits visual perception. PLoS One. 2013;8(9):e73629.

79. Jan JE, Groenveld M, Anderson DP. Photophobia and cortical visual impairment. DMCN. 1993;35(6):473–7.

80. Jan JE, Groenveld M, Sykanda AM, Hoyt CS. Behavioural characteristics of children with permanent cortical visual impairment. DMCN. 1987;29(5):571–6.

81. Dutton GN, Calvert J, Ibrahim H, Macdonald E, McCulloch DL, Macintyre-Béon C, et al., editors. Structured clinical history-taking for cognitive and perceptual visual dysfunction and for profound visual disabilities due to damage to the brain in children. 2010.

