## Supplemental Information for "Impaired reach-to-grasp integration identifies cerebral visual impairment (CVI) in children and adults"

**Supplementary Material**

*Exclusion Criteria*

Control participants were excluded from participating in the study if they: lacked stereopsis (depth perception) and/or had low vision due to eye misalignment or amblyopia (lazy-eye); had a new eyeglasses prescription in the last 2 weeks; were born prematurely at 34 weeks’ gestation or earlier; had any other notable eye impairments; had a diagnosis of cerebral palsy, epilepsy, hydrocephalus, or previously had meningitis; had any other condition that affects hand use, brain development, or vision; had any immediate family members diagnosed with a learning difficulty/disability, dyslexia or significant reading or writing difficulty, dyspraxia, developmental coordination disorder, ADHD, social communication disorder, schizophrenia, or any other specific language impairment; or had long fingernails that may interfere with the ability to complete the reach-to-grasp task.

Participants with CVI were recruited with the help of Dr. Isobel Hay, a paediatrician at the National Health Service (NHS) for Dumfries and Galloway in Scotland, UK. Inclusion criteria for participants with CVI were as follows: previously received a diagnosis of CVI from the CVI service (based on combined paediatric/orthoptic/ophthalmology assessments) at NHS Dumfries and Galloway in the absence of any ophthalmic condition or intellectual disability; previous neurological examination confirming no neuromuscular cause for their observed impairments; and profiles of functional impairment of vision were derived from previous completion of the Dutton CVI Inventory (81).

*Intellectual Property Ownership*

The reaching targets described in this publication (‘Target blocks for Optic Ataxia diagnosis/assessment’) contain material that is protected by United Kingdom copyright and trade secret law, and by international treaty provisions. All copyrights, patents, trade secrets, trademarks, service marks, trade names, moral rights and other intellectual property and proprietary rights in the (‘Target blocks for Optic Ataxia diagnosis/assessment’) shall remain the sole and exclusive property of NHS Dumfries & Galloway, Mountainhall Treatment Centre, Bankend Road, Dumfries, DG1 4AP, UK as applicable.
